# Impact of environmental temperature on Covid-19 spread: Model and analysis of measurements recorded during the second pandemic in Cyprus

**DOI:** 10.1101/2021.02.19.21252106

**Authors:** G. Livadiotis

## Abstract

The paper investigates the effect of the environmental temperature on the spread of COVID-19. We study the daily numbers of the cases infected and deaths caused by Covid-19 during the second wave of the pandemic within 2020, and how they were affected by the daily average-high temperature for the districts of the Republic of Cyprus. Among the findings of the paper, we show that (i) the average ratio of the PCR to rapid positive tests is ∼2.57±0.25, as expected from the tests’ responses, indicating that PCR overestimates positivity by ∼2.5 times; (ii) the average age of deaths caused by Covid-19 increases with rate about a year of age per week; (iii) the probability of a person infected by Covid-19 to develop severe symptoms leading to death is strongly depended on the person’s age, while the probability of having a death on the age of ∼67 or younger is less than 1/1000; (iv) the number of infected cases and deaths declined dramatically when the environmental temperature reaches and/or climbs above the critical temperature of *T*_C_=30.1±2.4 C^0^; (v) the observed negative correlation between the exponential growth rate of the infected cases and the environmental temperature can be described within the framework of chemical kinetics, with at least two competing reactions, the connection of the coronavirus towards the receptor and the dissolution of the coronavirus; the estimated activation energy difference corresponding to the competing chemical reactions, 0.212±0.25 eV, matches the known experimental value; and (vi) the infected cases will decline to zero, when the environmental temperature climbs above the critical temperature within the summery days of 2021, which is expected for the Republic of Cyprus by the 16^th^ of May, 2021.

## 1. Introduction

A recently published paper by PLOS ONE [1] showed the first reported statistically significant relationship of negative correlation between the average environmental (or weather) temperature and exponential growth rates of cases infected by Covid-19. This relationship led to the derivation of the critical temperature *T*_C_, for which the exponential growth rate becomes zero and thus the COVID-19 spread declines once the environmental temperature *T* climbs higher than this critical value. The critical temperature was estimated to be *T*_C_∼86.1 ± 4.3 F^0^ or *T*_C_∼30.1 ± 2.4 C^0^. The mentioned paper published first as a MedRxiv preprint in late April, 2020. Since then, several other studies across the globe also showed the negative correlation between the COVID-19 infected cases and environmental temperature or other meteorological parameters (e.g., see: [2-9]).

Here we study how the environmental temperature impacts the infected cases during the second wave of the pandemic. For this, we examine the number of the cases infected and the number of deaths caused by Covid-19, together with the daily average-high temperature of the infected cases for the districts of the Republic of Cyprus.

The first cases in Cyprus infected by Covid-19 confirmed on early March, 2020; this first wave of the pandemic peaked on early April,2020, with ∼40 daily new cases on average, and ended on the 23^rd^ of May 2020, the first day with zero new cases. The number of daily new cases were kept low, i.e., ∼1-3 on average, until mid-July, where they raised to 10-20 on average for about a month, before they dropped down again to ∼1-3 in early September, 2020. In late September, a persistent increase of the daily new cases was observed; this evolved to the second wave of the pandemic in Cyprus, peaked on the days from late December 2020 to early January 2021, reaching ∼600 daily new cases on average, and then decayed to about a hundred new cases on late January 2021 [10].

There is a significant difference between the method of recording the new cases in the first and second waves. During the first wave, people were tested in hospitals and other urgent care centers after feeling suspicious flu-related symptoms; therefore the majority of the confirmed cases were symptomatic. On the other hand, during the second wave of Covid-19 pandemic, confirmed cases or positive tests were counted with statistical sampling; therefore, the majority of the confirmed cases were asymptomatic, because only a small fraction of the infected population presents noticeable symptoms (see Section 2).

Once we understand the number of the infected cases against the number and type of the tests performed, as well as the involved statistical biases, we model the exponential growth phase of the time series of the cases infected and deaths caused by Covid-19, and compare them with the time series of the average-high environmental temperature. The arrangement of the paper is as follows: In Section 2, we present and investigate the recording of the infected cases during the second wave of the Covid-19 pandemic in the Republic of Cyprus. In particular, we (i) describe the data of the time series of environmental temperature and of daily numbers of cases infected and deaths caused by Covid-19, (ii) estimate the percentage of the asymptomatic infected cases, (iii) understand the statistical sampling performed and the involved biases, and (iv) investigate the types of tests and their reliability. In Section 3, we model the evolution within the exponential growth phase of the cases infected by Covid-19. In Section 4, we analyze the number of deaths caused by Covid-19 and their age distribution. In Section 5, we investigate the impact of environmental temperature on the numbers of the cases infected and deaths caused by Covid-19. In Section 6, a theoretical explanation of the inverse-temperature dependence detected, is explained within the framework of chemical kinetics. In the discussion of Section 7, we forecast the decline of the number of infected cases when the environmental temperature will climb above this critical value within the summer of 2021 for the Republic of Cyprus. Finally, Section 8 summarizes the conclusions.

## 2. Recording the infected cases during the second wave of the Covid-19 pandemic

### 2.1. Data

We use publicly available datasets of (1) the environmental average-high temperature (e.g., see: [11]); (2) time series of the daily number of the infected cases and the deaths caused by Covid-19 (e.g., see: [10]) for the districts of the Republic of Cyprus.

### 2.2. Percentage of symptomatic cases

During the second wave of the Covid-19 pandemic, confirmed cases or positive tests were counted with statistical sampling; therefore the majority of the confirmed cases were asymptomatic, because only a small fraction of the infected population presents symptoms, at least significant ones to be taken into account. In order to understand better this, we examine the official daily data sets as announced by the government of Cyprus [10]. As an example, we read the announcement of the 24^th^ of October, 2020, translated in Table 1.

**Table 1.**
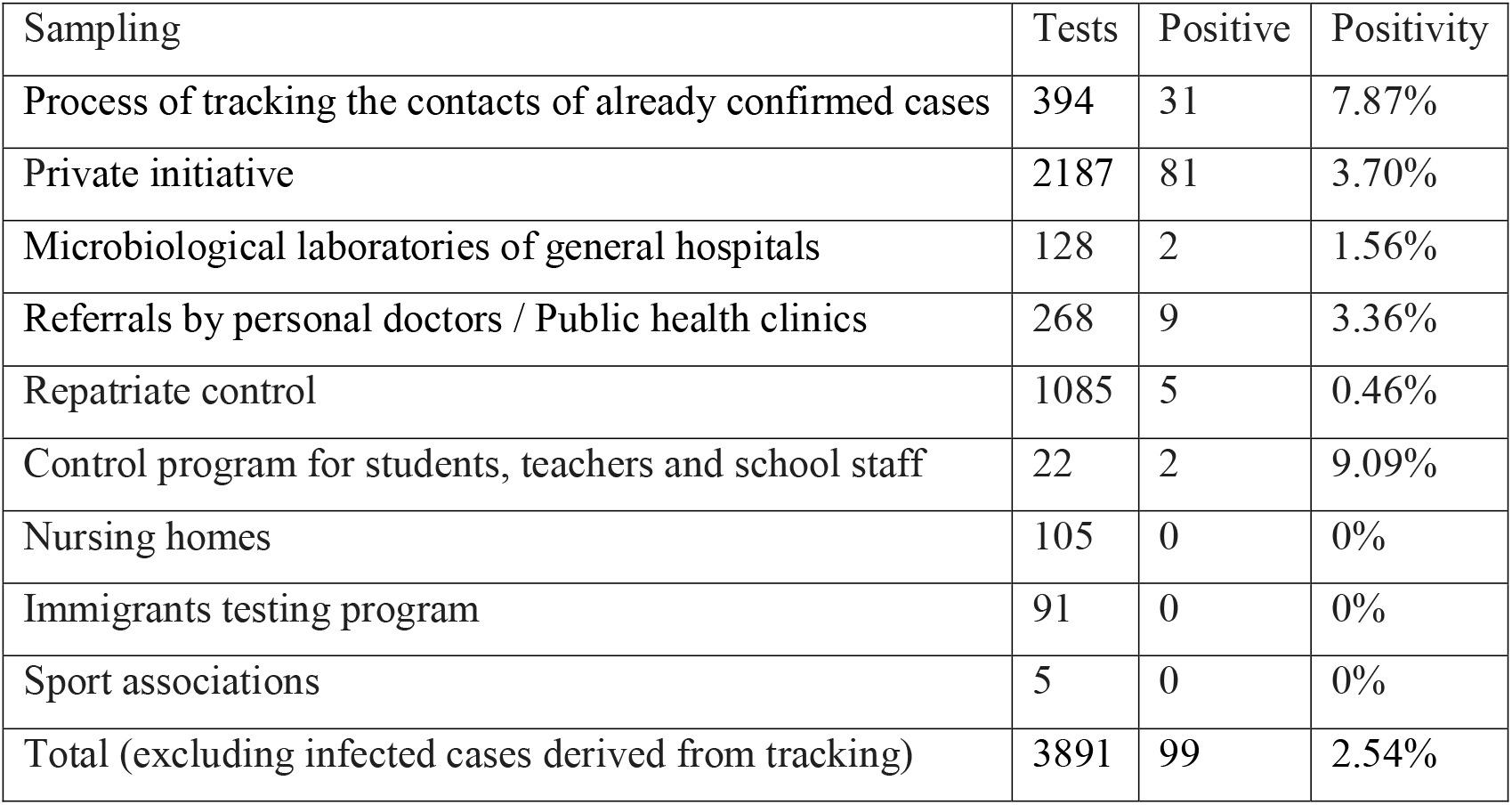
Analysis of the confirmed cases and performed cases on the 24^th^ of October, 2020 [15]

According to Table 1, there were 2583 tests taken place in hospitals, health clinics, or in laboratories from private initiatives, that is, 2583 people felt like they have some sort of worrying symptoms, from which 92 cases were confirmed to be infected by Covid-19. Also, there were 7 infected (mostly asymptomatic) cases out of 1308 tests sampled from diverse groups. Therefore, at that certain single day, and for the whole country, there were confirmed 92 symptomatic infected cases, i.e., people who felt symptoms and thus they were concerned enough to visit hospitals, clinics, or laboratories for testing, which came back positive for Covid-19. On the other hand, there were 7 asymptomatic infected cases out of 1308 samples, corresponding to ∼6500 asymptotic cases for the whole population of Cyprus Republic (∼1.2 millions as of February 2021 [12]). Overall, there were 92 symptomatic cases (with symptoms intense enough to have people concerned about them and have tested), from which 11 cases were probably severe enough to be tested in a hospital/clinic), and an estimation of ∼6500 asymptomatic cases; therefore, the symptomatic cases are ∼1.4%. If we assume that from the 81 cases confirmed from tests performed after private initiative, there were *x* asymptotic cases and 81−*x* symptomatic ones, then the estimation for the whole population is ∼1.2·10^6^ (7+*x*)/(1308+2187) asymptotic and 11+81−*x* symptomatic, corresponding to a percentage of symptomatic cases that maximizes for *x*→1 (at least one asymptotic case), i.e., ∼ 3.2%. For one incubation period, τ ≈ 5.2±1.1 d [13], we have about the same number of asymptomatic cases, but the symptomatic cases should be counted for each day of the period τ, i.e., ∼ (11+81−*x*)·τ; thus, the maximum percentage of the symptomatic cases is ∼14.3%, while if *x*=4 asymptomatic cases exists within the 81 confirmed cases, the percentage drops to ∼10.5%.

Note that the key-point is to understand that the hospital/clinics and private initiate’s related cases are not considered random statistical sampling and they are representative for the whole population (i.e., without having to estimate the numbers for ∼1.2·10^6^). Then, we estimate that for the Cyprus population, and at this date, the symptomatic cases constitute about one tenth the confirmed cases, or less. It has to be clarified though that the determined asymptomatic cases include also cases with minor symptoms which have been disregarded. The absolute zero symptoms cases may be about the same order as the symptomatic cases [14].

### 2.3. Statistical sampling and biases

The record of the infected cases during the second wave of the Covid-19 was based on a daily statistical sampling of the population. The positive tests constituted the confirmed infected cases within the statistical sampling. The ratio of the positive to the total number of tests is called positivity percentage, and for estimating the infected cases for the whole population one has to multiply the positivity with ∼1.2·10^6^.

The performed sampling was biased because of the following reasons:

- The random sampling was taking into account the number cases confirmed by tracing; however these cases were included within the personal circle of friends/relatives of previously confirmed cases.
- The random sampling was also counting the tests performed in hospitals/clinics and those performed by private initiative, which are representative of the whole population, instead of the selected random sample.
- The number of tests was not constant, characterized by large variations, ranging from less than a hundred to a few ten thousands.

In Figure 1 we plot the positivity percentage for tests performed in the five districts of the Greek Cyprus (Nicosia, Limassol, Larnaca, Paphos, Famagusta), normalized by the Country’s average positivity percentage. We observe less variability as the number of the Country’s average ratio of infected cases increases.

**Figure 1.**
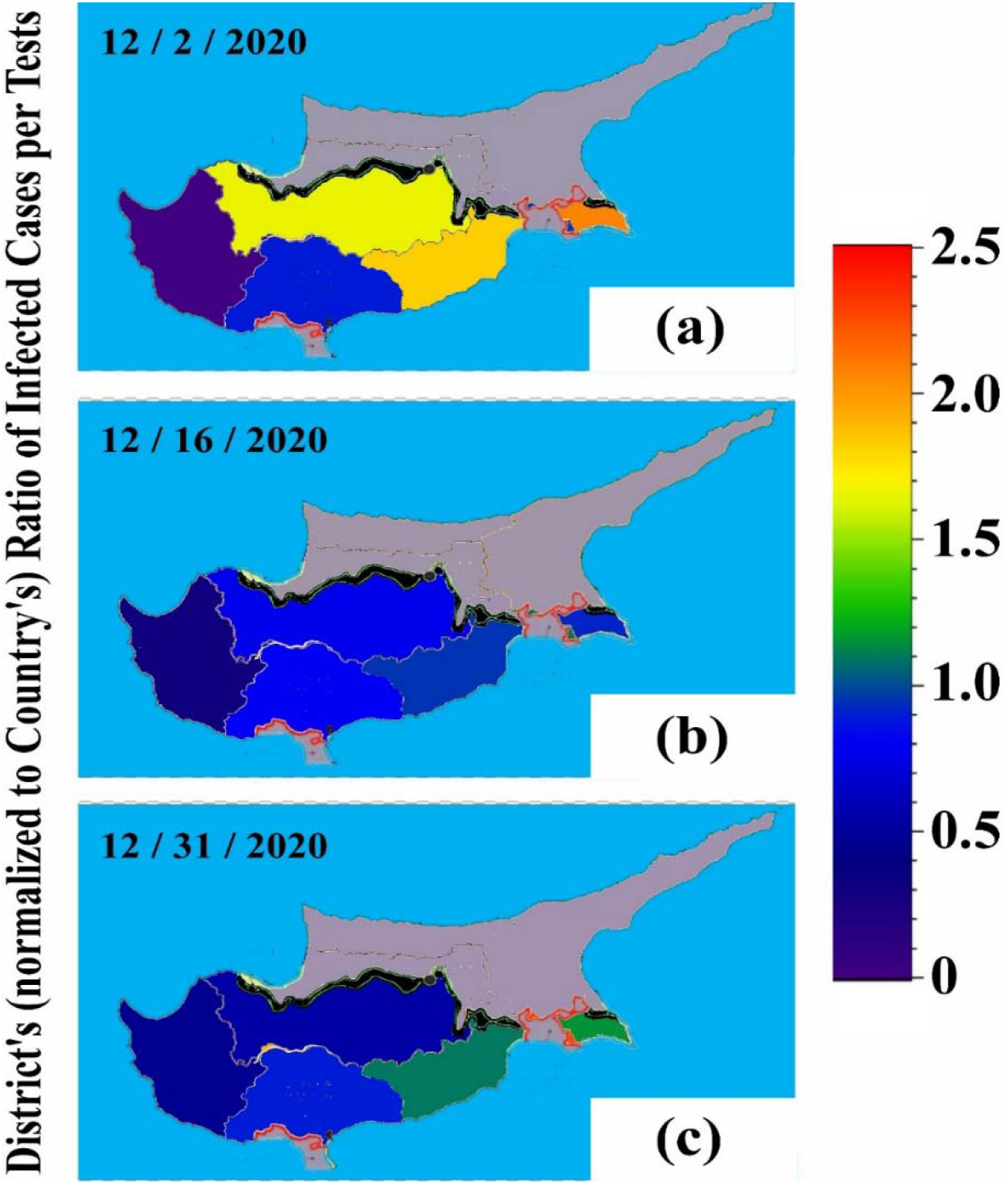
Variability of the positivity percentage, i.e., the ratio of the infected cases per total number of tests, for sampling performed in the five districts of Greek Cyprus (Nicosia, Limassol, Larnaca, Paphos, Famagusta), normalized by the Country’s average respective ratio. (Note: The occupied north part of the island is grey colored.)

### 2.4. Types of tests and reliability

Starting on the 16^th^ of November, both PCR and rapid antigen tests were performed in the Republic of Cyprus. Positive rapid tests had to be confirmed with a PCR test until Christmas, 2020. However, it was soon realized that the rapid tests had been almost always confirmed by PCR tests (∼98% success); therefore, beyond 12/25/2000, PCR tests were not required for confirming the infection.

The rate of detecting infections of antigen tests is between 84% and 98% (if a person is tested in the week after showing symptoms). The probability of detecting Covid-19 infection differs for rapid antigen and PCR tests. For both the tests, the probability starts to be nonzero as soon as the exposure period ends. For the rapid test, the probability peaks in one week and becomes negligible in about two weeks (∼17 days) from the exposure period; for the PCR, the probability peaks in two weeks and starts to tend to zero in about five weeks from the exposure period. The PCR’s response is characterized by a nonzero probability for more than five weeks, that is, ∼ 5 / 2 = 2.5 times compared to the rapid test [16].

Figure 2(a) plots the ratio of the positive PCR to the positive rapid tests (time series and distribution). The mean is estimated to be 2.57 ± 0.25, that is, the expected value, as explained above. In Figure 2(b) the distribution of the ratios was constructed by generating 1000 points for each point of the original time series, using the method of population magnification. In particular, for each of the given *N*_0_ data points of 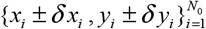, we reproduce *N*=1000 bi-normally distributed data points, ∼ *N* {*μ*_*x*_≡*x*_*i*_,*σ*_*x*_≡*δx*_*i*_} × *N* {*μ*_*y*_≡*y*_*i*_,*σ*_*y*_≡*δy*_*i*_}; (for further details, see: [17]). The mode of the distribution coincides with the mean value derived in Figure 2(a); in addition, a Normal distribution is fitted with mean and error given by 2.60 ± 0.26.

**Figure 2.**
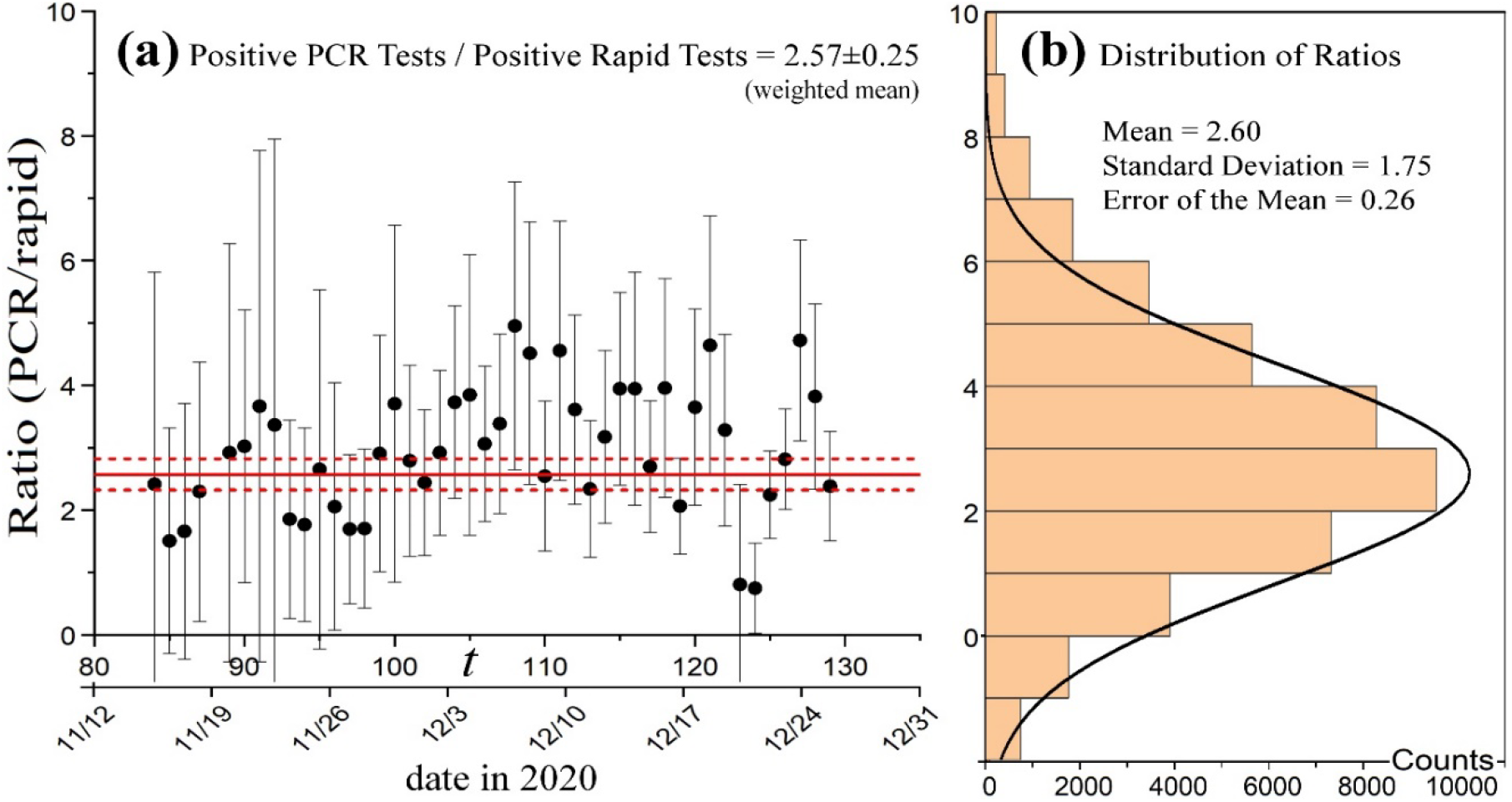
(a) Time series and (b) distribution of the ratios of the detected infected cases per PCR tests, divided by the detected infected cases per rapid antigen tests.

We observe that the average ratio of the PCR positive tests to the rapid positive tests, as performed in Cyprus in 2020, is given by 2.57 ± 0.25; this coincides with the expected ratio of the PCR’s response to the rapid test response.

Early research suggested that it takes 2 weeks for the body to get over a mild illness. On the other hand, data indicate that persons with mild to moderate COVID-19 remain infectious no longer than 10 days after symptom onset [18]. Therefore, a “positive” PCR result reflects only the detection of viral RNA and does not necessarily indicate presence of viable virus [19]. PCR’s response is characterized by a nonzero probability for more than 5 weeks; hence, it provides an overestimation of the positive tests, and thus, of the infected cases, about ∼5/2 = 2.5 times compared to the rapid test [16].

It is generally expected that a response with only two weeks of nonzero probability is satisfactory and reliable for detecting the infection. The PCR’s response is characterized by a nonzero probability for more than 5 weeks; hence, it provides an overestimation of the positive tests, and thus, of the infected cases (about 2.5 times compared to the rapid test) [18].

## 3. Modeling of the evolution of cases infected by Covid-19

The basic model for describing the evolution of the infected cases is based on the combination of two factors [20]: (i) the per-capita growth of its population, represented by a function proportional to the population, e.g., *E*(*x*) = λ. *x*, and (ii) the negative feedback that models the factors that flattens the curve, e.g., *I* (*x*) = 1 − *x*^*b*^,

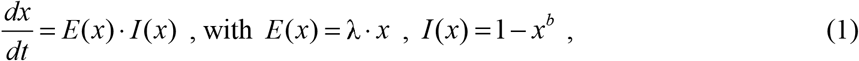

Where *x*(*t*) ≡ *N* (*t*) / *N*_max_, *x*_0_ ≡ *N*_0_ / *N*_max_ ; *N*(*t*) is the number of total infected cases evolved from the initial *N*_0_≡*N*(0) cases, *N*_max_ is the maximum possible number of infected cases.

The function of negative feedback models the factors that flattens the curve, such as, the measures taken against spreading. These factors are not affecting the exponential growth rate λ, but they do flatten the curve in the presence of the exponential growth. In other words, the exponent *b* controls the effectiveness of the factors that flattens the curve, but it is not affecting the value of λ itself or the function *E*(*x*). Indeed, strict {loose} measures correspond to smaller {larger} values of *b*.

We are interested in studying the physical mechanisms behind the value of exponential growth rate λ. For this, we hereafter ignore the value of *b*, setting it at zero and considering only the exponential growth modeling, i.e.,

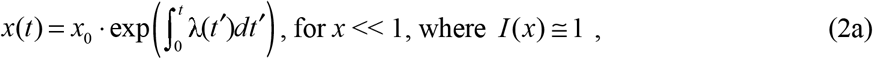

that is,

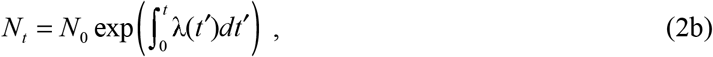

or, for slowly changing rate,

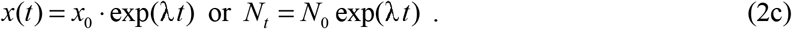

The exponential rate is given by [1,21]:

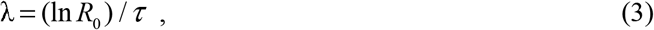

where τ is the incubation period [13], and the reproduction number *R*_0_ is defined as the average number of people that a single infected person will spread the disease [22], that is, a measure of how contagious a disease is, and depends on the physical characteristics of coronavirus [23]; Characteristic values for COVID-19 is *R*_0_∼2-4 [13].

The main factors that can affect the exponential rate λ are: (a) culture in social activities, and (b) environmental temperature and/or other thermodynamic parameters [1-9]. Intense cultural and social activities have frequently close contacts, leading reasonably to a positive correlation with *R*_0_. Measures against the virus spread prevent the exponential growth leading soon or later to a decay, but do not modify the exponential rate which is a characteristic of the population and its thermodynamics with environment and weather.

The detailed statistical analysis of data sets (taken from Italy and USA), performed in [1], showed that the rate can be modeled by (i) a parameter *p*_1_ affected by the culture of the population, and (ii) a parameter *p*_2_ primarily affected by the environmental temperature; namely,

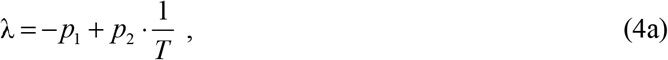

rewritten as

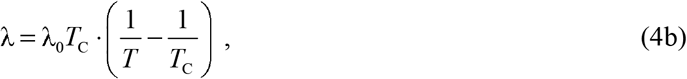

where the critical temperature, for which the exponential rate becomes zero ceasing the exponential growth, is given by [1]:

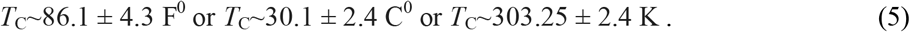

Next we model the exponential growth of the infected cases per number of performed tests (positivity percentage), separately for the PCR and rapid tests. There is a slow variation of the average daily environmental temperature; this can be empirically modelled by

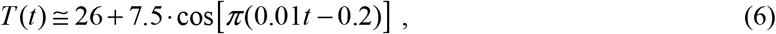

where time *t* is in days, with the day *t*=0 corresponding to the 8/25/2020. Then, we substitute Eq.(6) in the following to derive the affected cases to model the number of the infected cases, *N*_*t*_,

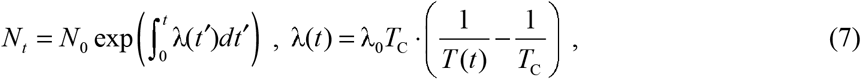

which has to be divided by the total population ∼1.2·10^6^. Figure 3 plots the ratios of the observed infected cases per number of PCR (red) tests or total (PCR and rapid) tests (blue), recorded from 8/25/2020 to 12/31/2020. The time series interval corresponding to the exponential growth is modelled by Eq.(7) (black).

**Figure 3.**
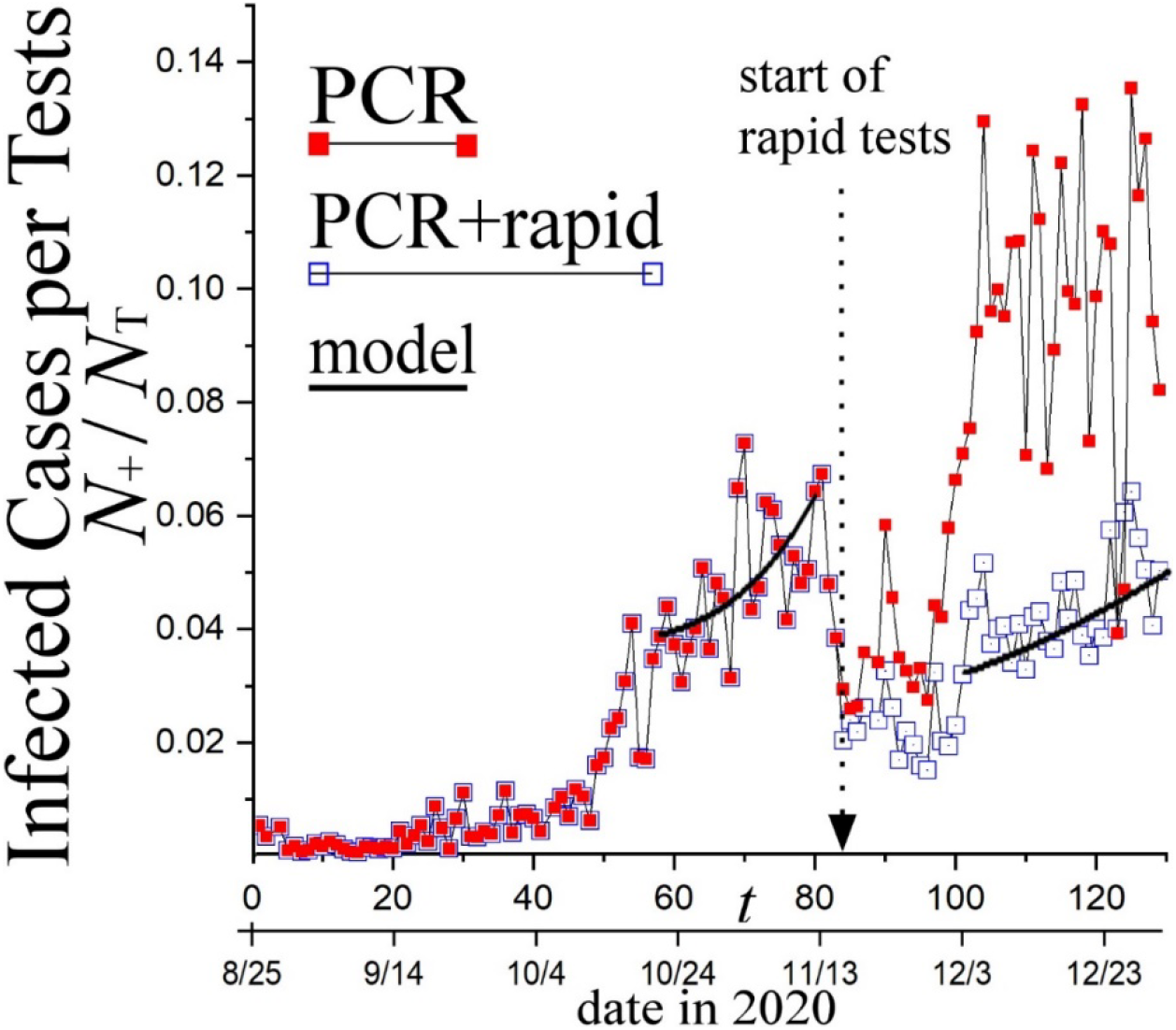
Time series of the positivity percentages, i.e., the ratios of the infected cases per PCR (red) tests or both (PCR and rapid) tests (blue); the latter is calculated as the total number of positive tests divided by the total number of tests. The model in Eq.(7) shows the exponential growth rate (black). The parameter values used are λ_0_ *T* = (2800 K)/τ, *N*_0_ = 10^5^ and (640 K)/τ, *N*_0_ = 0.32·10^5^, respectively for PCR and all tests.

## 4. Analysis of the age of deaths caused by Covid-19

The vast majority of the recorded deaths caused by Covid-19 were with underlying health conditions. Interestingly, the moving average increases with time. Figure 4 plots the mean and its error of the age of deaths for moving windows of (a) 5 days and (b) 10 days. The detected trend is:

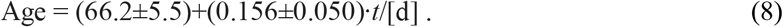

**Figure 4.**
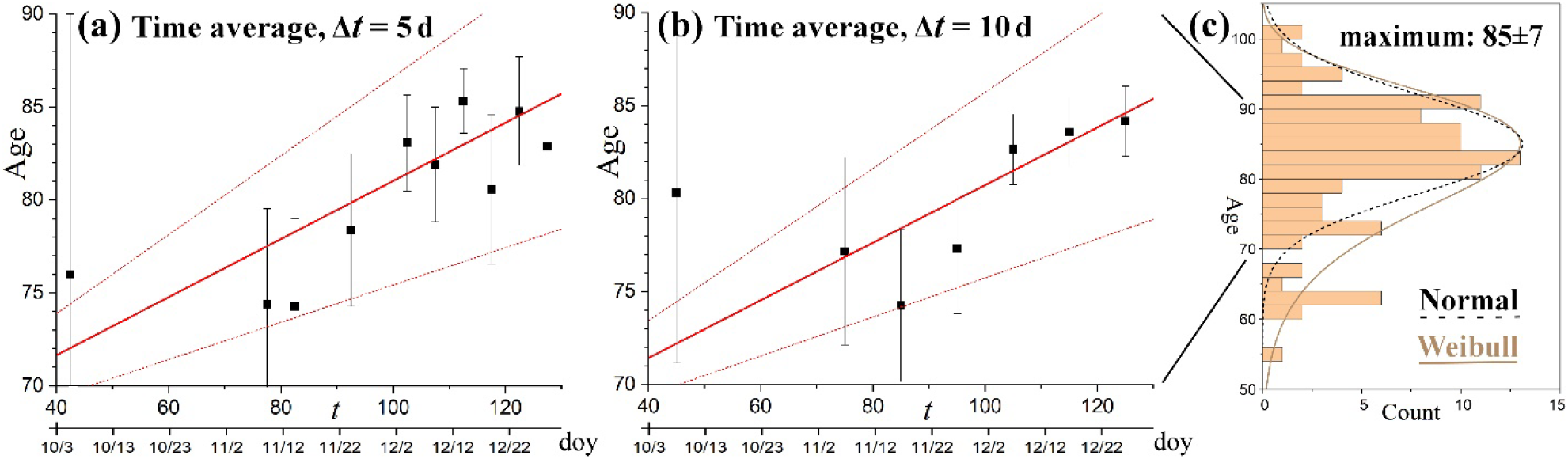
Smooth-averaged time series of the age of the deaths by Covid-19 for time window for averaging (a) Δ*t*=5d and (b) Δ*t*=10d. (c) The distribution of the age of all deaths by Covid-19; the fitted curves show the Weibull and Normal distributions.

The last ∼10 days of 2020 the average age appears to be approaching the mode of the whole age distribution (plotted in Figure 4(c)), that is ∼85±7.

The age distribution of the deaths, *N*_d_·*P*(Age), plotted in Figure 4(c), is fitted by the Normal distribution *P*(Age) with mean *μ* = 85 and standard deviation *σ* = 7. This distribution has to be weighted by the age distribution of the population in the Republic of Cyprus [24], which can be also fitted by a Normal distribution with *μ* = 34 and *σ* =25. The weighted distribution shifts the age distribution to larger ages with *μ* = 89.3 and σ = 7.2 (see Figure 5).

**Figure 5.**
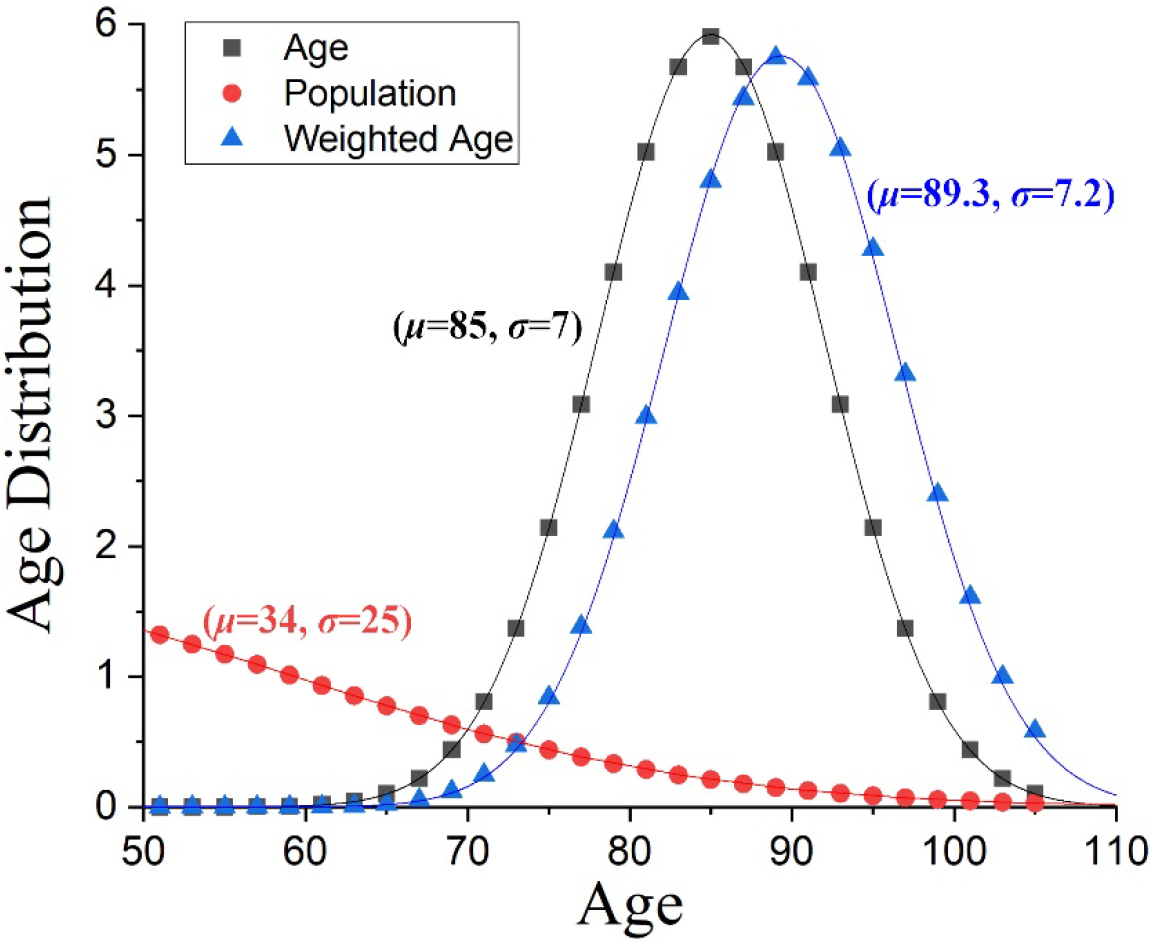
Age distribution of the deaths caused by Covid-19, *N*_d_·*P*(Age), plotted for *N*_d_=104, and *P*(Age) is the Normal distribution with (*μ*=85,*σ*=7) for the best fit in Figure 4(c) (black). When weighted by the age distribution of the population in Cyprus (red), the resulting weighted age distribution becomes to be expressed by the Normal distribution with (*μ*=89.3,*σ*=7.2) (blue).

Figure 6(a) plots the cumulative Normal distribution with mean *μ*=89.3 and standard deviation *σ*=7.2, which constitutes the weighted Normal distribution of the number of deaths in Figure 5. This is given by

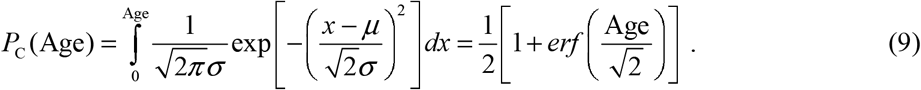

**Figure 6.**
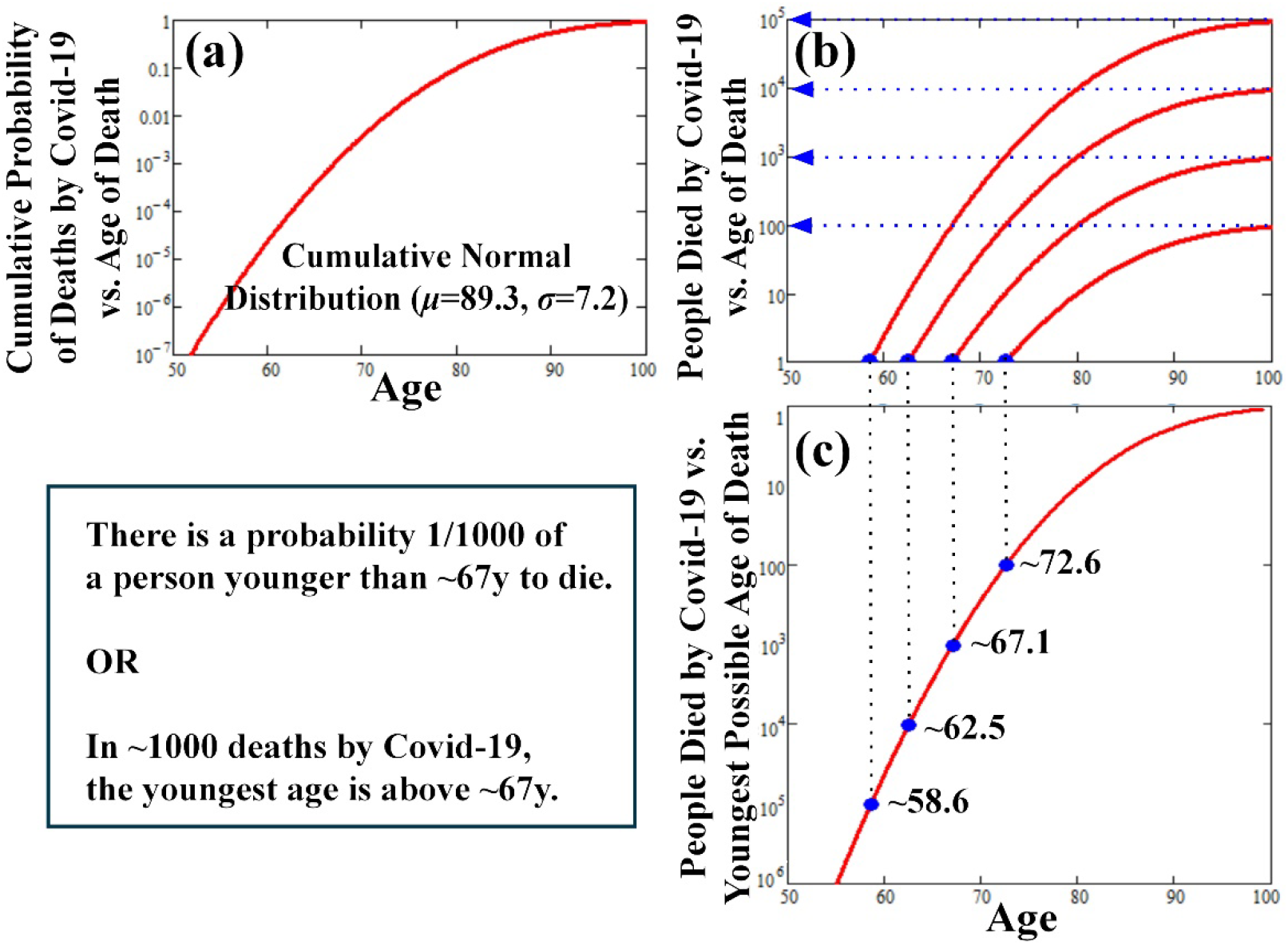
(a) Cumulative Normal distribution with mean *μ*=89.3 and standard deviation *σ*=7.2, representing the age of deaths by Covid-19, *P*_C_(Age). (b) Cumulative age distribution of a number of deaths, *N*_d_·*P*_C_(Age). (c) Number of the deaths by Covid-19, *N*_d_, plotted as a function of the youngest age (Age); this is the number of the deaths *N*_d_(Age) which is expected before we observe a death with age given by “Age”.

The age cumulative probability distribution, multiplied by the number of total deaths *N*_d_, determines at what age we expect to have *N*_d_=1. In Figure 6(b) plots *N*_d_·*P*_C_(Age) for *N*_d_ = 10^*γ*^ with *γ*=2, 3, 4, and 5. Therefore, the age for which the number of deaths becomes less than one is, respectively, 72.6, 67.1, 62.5, and 58.6 (Figure 6(c)). According to the same cumulative probability distribution, there is a probability 1:1000 of a person younger than ∼67 y.o. to have died from Covid-19 in the examined time interval.

The conclusion that can be interpreted from Figure 6 is the following: the probability of a person infected by Covid-19 to develop severe symptoms leading to death is strongly depended on the age of the person. In particular, the probability of having a death on the age of 67 or younger is less than 1/1000, or, among 1000 deaths by Covid-19 there would be only one case expected with age ∼67. Even if all the daily asymptomatic cases of the whole population (10^*γ*^, with *γ* = 4 - 5) were suddenly shown severe symptoms leading to death, the youngest age is expected to be above ∼62. (This would require about 8% of the population to be above the age of 55; but the population in Cyprus is old enough as 15% of the population ages are above 65 [24].)

## 5. Impact of environmental temperature

Here we investigate the impact of the environmental temperature on the number of daily new cases infected and the number of deaths caused by Covid-19. In [1] it was shown that rate of the exponential growth of the cases infected by Covid-19 decreases when temperature increases, while it is reduced to zero when the temperature reaches, or escalates above, the critical temperature of *T*_C_∼86.1 ± 4.3 F^0^ or *T*_C_∼30.1 ± 2.4 C^0^.

Figure 7 plots from 8/25/20 to 12/31/2020: (a) the daily infected cases normalized by the number of tests (positivity percentage) – as in Figure 3, (b) the daily environmental average-high temperature, and (c) the number of deaths by Covid-19 on a log-scale.

**Figure 7.**
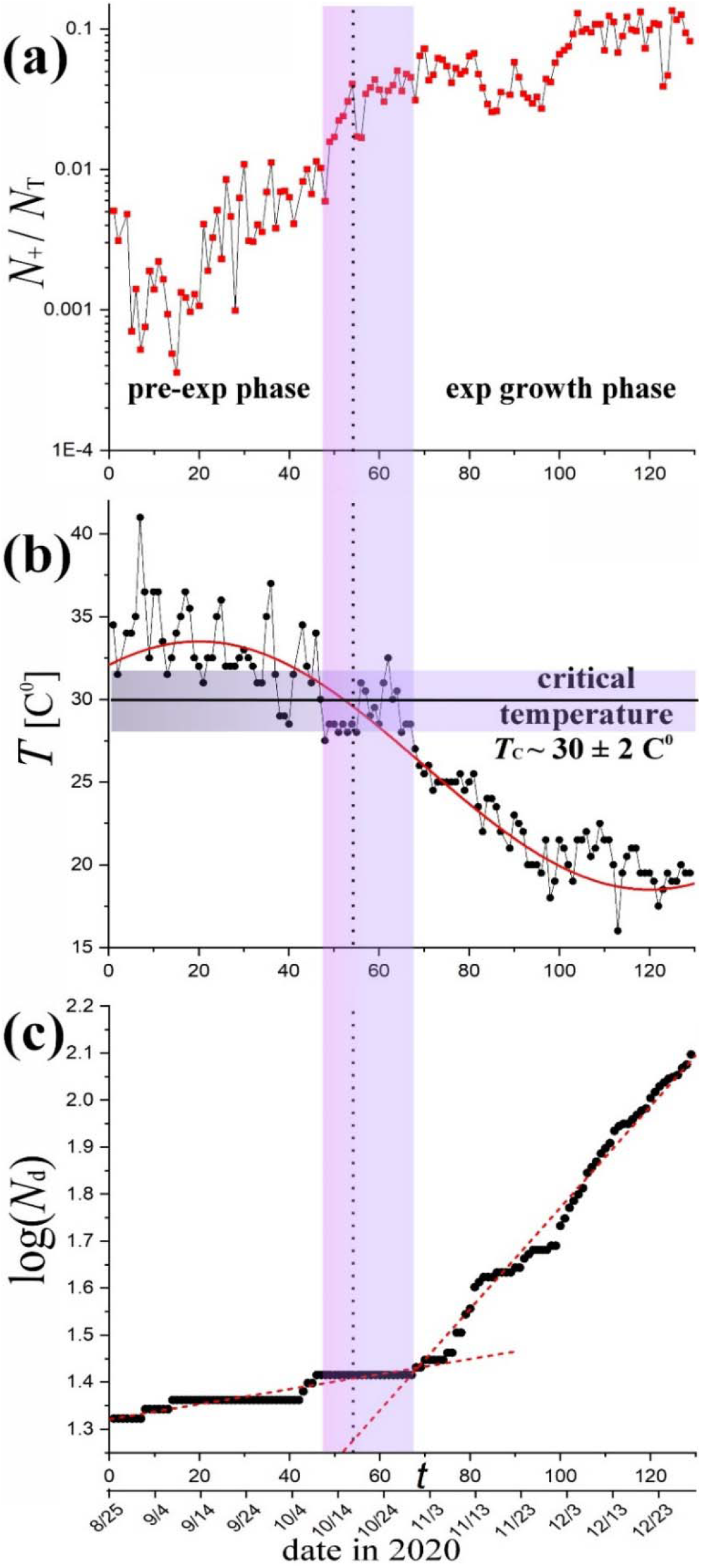
(a) Daily infected cases normalized by the number of tests (defined as positivity percentage); there are two subsequent phases plotted, the pre-exponential (pre-exp) and exponential (exp) growth phases, which had occurred before the number of the cases started to decline; (b) Daily average-high environmental temperature (also averaged over the districts of Nicosia and Limassol), and (c) the number of deaths by Covid-19 on a log-scale; all three panels have their horizontal axis aligned, indicating the time in days of the pandemic second wave during 2020, spanning from 8/25 to 12/31.

We observe that by mid-October and thereafter, the temperature dropped below its critical value *T*_C_. A transition region appears between mid and end of October. The (normalized) number of infected cases, recorded with PCR tests after mid-October, increased and escalated up to one order-of-magnitude higher than the respective number of cases recorded before mid-October. (Note: the infected cases recorded with rapid test is excluded from the analysis with respect to the environmental temperature. The number of the daily infected cases recorded with rapid tests is about 2.5 times lower than that of PCR; as shown and discussed in Subsection 2.4, this is caused by an overestimation that likely characterizes PCR’s response, that is, of about 2.5 times compared to the rapid test response.)

In addition, we observe that the number of deaths increased rapidly after mid-October. The rate of log(*N*_d_), where *N*_d_ is the number of the daily new number of deaths, is significantly smaller for *t* < 10/19/2020, when *T* > *T*_C_, than for *t* > 10/19/2020, when *T* < *T*_C_. In order to show this, we perform a statistical analysis to fit the two-parameter linear statistical model *y*(*x* ; *p*1, *p*_2_) = *p*_1_ + *p*_2_ *x* to the given *N* data points, where *x* ≡ *t, y* ≡ log(*N*_d_). Therefore, we minimize the total square deviations, or the chi-square for a unit standard deviation,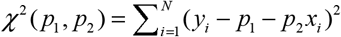. The global minimum of the chi-square function *χ* ^2^ (*p*_*1*_, *p*_*2*_) gives the optimal parameter values, 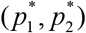, by solving the normal equations ∂*χ* ^2^ (*p*_*1*_, *p*_*2*_) / ∂*p*_*1*_ = 0 and ∂*χ* ^2^ (*p*_*1*_, *p*_*2*_) / ∂*p2* = 0 ; the minimum chi-square value is 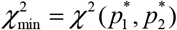. The statistical errors of these values are given by 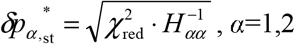, where *H* is the Hessian matrix of the chi-square at the global minimum, and 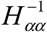 is the *α*-th diagonal element of its inverse matrix [25-27];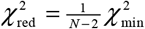 is the reduced chi-square value for *N*-2 degrees of freedom.

The linear fit, before and after mid-October is, respectively, given by

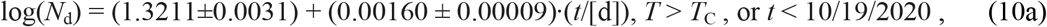

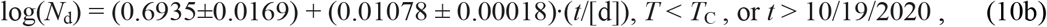

as we can see, the rate of the logarithm of the number of deaths increased by ∼6.7 times after the mid-October, that is, when the temperature dropped below the ∼ 30 C^0^.

Next we examine the number of the infected cases directly against the environmental temperature. In particular, in Figure 8, the confirmed infected cases *N*_+_ (positive tests), normalized to the number of the PCR tests *N*_T_, i.e., the percentage of positivity, is plotted as a function of the environmental temperature *T*. We observe that the normalized infected cases drops down by 3-5 times as the temperature values increase from ∼15 C^0^ towards the critical temperature *T*_C_; however, when the temperature reaches and exceeds its critical value, the infected cases decreased with much steeper rate (i.e., ∼30 times larger rate - absolute value). In the linear scale in Figure 8(a), the infected cases appear to drop down to zero, while their steep decreasing rate is clearer in the log scale shown in Figure 8(b); the infected cases per tests decreases by about two orders of magnitude just within the ribbon of temperatures 30 ± 2 C^0^.

**Figure 8.**
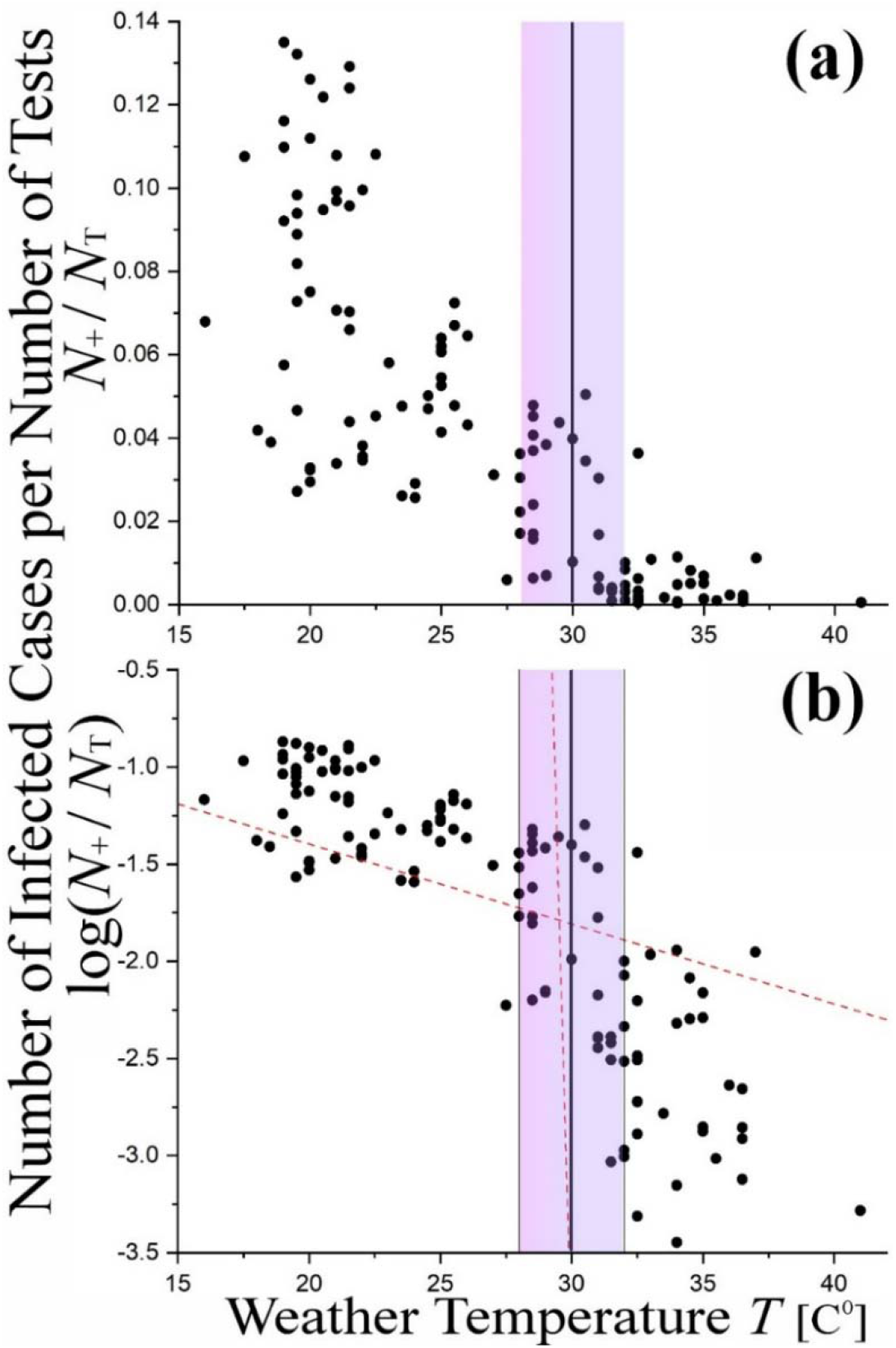
The percentage of positivity, i.e., the number of the detected infected cases (positive tests) per the number of the PCR tests, is plotted as a function of the weather temperature in both linear (a) and logarithmic (b) panels. In (a) the infected cases appear clearly to decrease with increasing temperature and vanish as soon as the temperature reaches ∼ 31 C^0^, that is, the limit of critical temperature *T*_C_ = 30 ± 2 C^0^ (purple ribbon). In (b) we observe two different functional behavior on each side of the ribbon; the number of infected cases decreases with much steeper slope for *T*>*T*_C_ than for *T*<*T*_C_; the corresponding expressions are given in Eqs.(11a,b), and plotted with −1*σ* (red dash lines) to show the temperature (∼29.5 C^0^) at which the two data regions intersect.

We now perform the fitting of *y*(*x* ; *p*_1_, *p*_2_) = *p*_1_ + *p*_2_ *x* with *x* ≡ *t, y* ≡ log(*N*_+_*/N*_T_). Therefore, we minimize the chi-square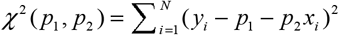. The fitting of the two trends (e.g., [25-27]) leads to

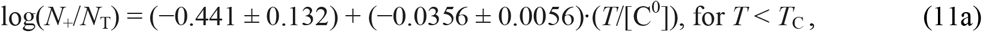

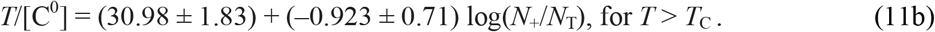

Next, we perform a statistical analysis to examine whether the determined temperature dependence of the infected cases and deaths is subject to statistically significant concentrations or rarefactions; namely, whether possible heterogeneities within the distribution of sample points plays significant role in the fitted relationships in Eqs.(10a,b) and (11a,b). For this, we derive the temperature infected relationship by fitting the homogenized set of sample points, instead of the raw sample points; then, we examine whether the fitting parameters differ from those derived from fitting the raw sample points.

We homogenize the sample points by grouping them in temperature binning of Δ*T*=2C^0^ (e.g., see similar analyses in [28,29]). Then, we estimate the weighted mean and error of the rates included in each bin. Figure 9(a) shows the homogenized dataset and its fitting.

**Figure 9.**
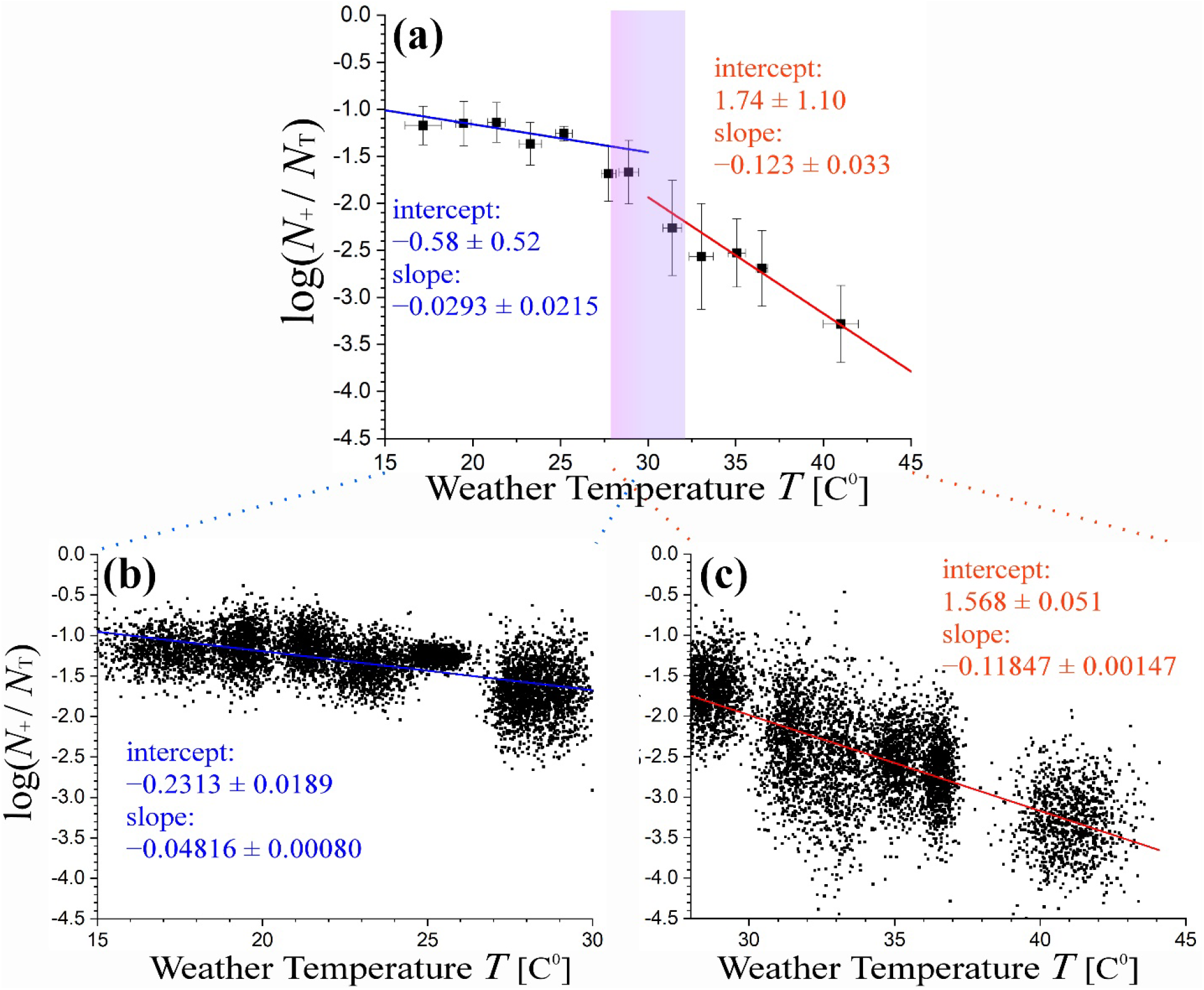
(a) Linear fitting of the (logarithm of) positivity percentages with respect to the weather temperature – similar to Figure 8(b), but with the homogenized dataset. (b) The fitting analysis is repeated for the “enriched” dataset (see text), in order to ensure that the statistical significance of the difference of slopes *d*log(*N*_+_/*N*_T_)/*dT*, taken for *T*>*T*_C_ than for *T*<*T*_C_.

We observed that homogenized datasets result in a smooth relationships between the values of binned temperature and the (logarithm of) positivity percentages. The linear relationship for *T*<*T*_C_ has large slope *d*log(*N*_+_/*N*_T_)/*dT* characterized by high statistical confidence for the raw or homogenized datasets (p-values much higher than the significant limit of 0.05). Therefore, the arrangement of sample points do not affect significantly the fitting results. We confirm this result by applying the method of generating data points, as in Figure 2(b), and then, refit the “enriched” dataset. In particular, for each of the given *N*_0_ = 12 data points of 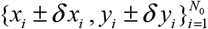, with *x* ≡ *T* and *y* ≡ log(*N*_+_/*N*_T_), we reproduce *N*=1000 bi-normally distributed data points, ∼ N {*μ*_*x*_≡*x*_*i*_,*σ*_*x*_≡*δx*_*i*_} × N {*μ*_*y*_≡*y*_*i*_,*σ*_*y*_≡*δy*_*i*_}; (for further details, see: [17]). The results are shown in Figure 9(b,c), confirming the statistical significance of the difference of slopes *d*log(*N*_+_/*N*_T_)/*dT*, taken for *T*>*T*_C_ than for *T*<*T*_C_.

## 6. Model of temperature dependence: Chemical kinetics

As it has already been discussed in [1], the observed dependence on temperature on the number of the infected cases and death caused by Covid-19 can be interpreted within the framework of chemical kinetics.

Coronavirus uses their major surface spike protein to bind on the ACE2 receptor — another protein that acts like a doorway into a human cell [30]. As long as the exponential growth takes place, the environmental temperature has an effective role on the chemical reaction between virus and spike protein. The whole process is a slow chemical reaction, where the mechanism behind can lead to rates negatively correlated with temperature, i.e., increasing rate with decreasing temperature. This is consistent to reaction rate proportional to the Arrhenius exponential with negative activation energy exp [| *ε*_*a*_ | / (*k*_B_*T*)] [1,31].

We recall that the exponential growth phase, which starts at some time *t*=*t*_0_ following exponential phase characterized by a mild logarithmic and often vague growth, is given by

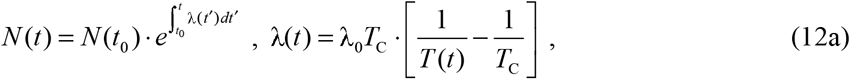

or, for a constant exponential rate,

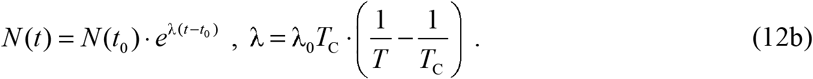

A physical interpretation of the inverse proportionality of the exponential growth rate with the environmental temperature could be given through the kinetics of the chemical reaction between the coronavirus (C) and the receptor (A),

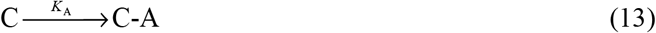

Or

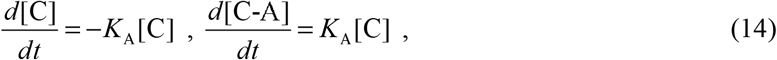

where the coefficient of the reaction speed is

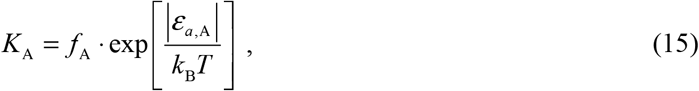

that involves the pre-exponential Arrhenius frequency factor *f*_A_ and the (negative) activation energy ε_*a*,A_. This is leading to 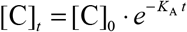, and using Eq.(14), we find

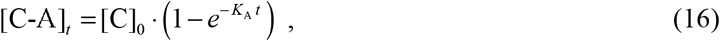

After one incubation period, τ, the concentration of the virus binding the receptor is [C-A]_*τ*_ ; the initial concentration of virus led to the final concentration of the infected receptor [C-A]_*τ*_, hence, the reproduction number *R*_0_ is

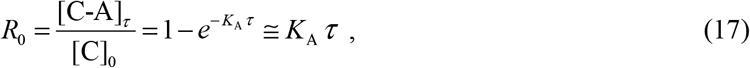

and the expansion rate becomes

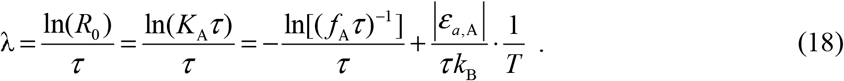

The quantity *f*_A_*τ* reads the probability of a collision between a coronavirus and a receptor at infinity temperature; then, (*f* _A_*τ*)^−1^ > 0, thus Eq.(18) has the same scheme as the exponential rate in Eq.(12b), as long as the activation energy is negative. In this case, we obtain

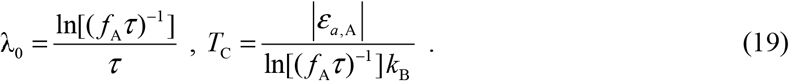

In a more realistic model, the coronavirus is not always reaching the receptor, because it is being destroyed in the air (product denoted by C-O), or dissolved through a reaction that competes the one that describes the connection with the receptor. The combination of both the parallel reactions can lead to the inversely-proportional dependence of λ on temperature. Indeed, we have

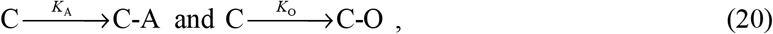

or

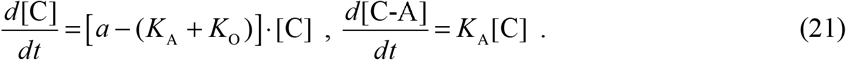

This is leading to 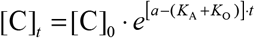, and using Eq.(21), we find

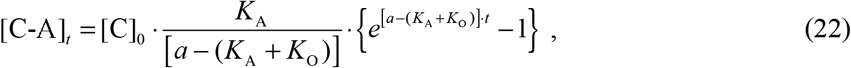

or

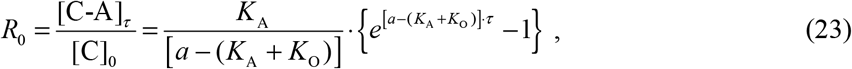

which can be approximated, as in the previous model in Eq.(17), by assuming sufficiently slow reactions,

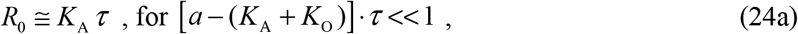

leading to the same results as in Eq.(19). On the contrary, in the approximation of fast reaction, Eq.(23) gives

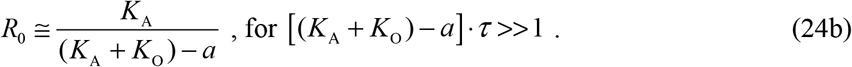

Then,

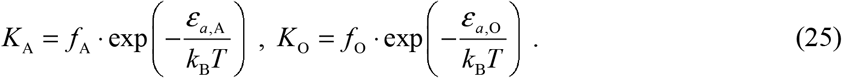

Next, we apply the following approximations: (i) ignore any virus-related source, i.e., *a* / *f*_A_ << 1; (ii) consider that the collisions of the coronavirus with the receptor, localized in the body’s upper respiratory systems, are much rarer with the collisions of the coronavirus with any other (air) particles causing its dissolution, i.e., *f*_O_ / *f*_A_ >> 1; and (iii) assume that a single infected person spreads the disease to *n* other persons per day and for (at least) τ days. Hence, the ratio of the total concentration of the infected receptors out of the initial concentration of coronavirus, *R*_0_, gives

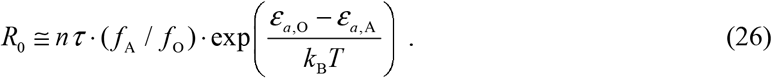

Hence, the exponential rate is given by

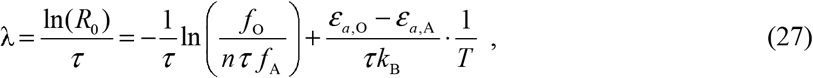

thus, comparing to the exponential rate in Eq.(12b), we find

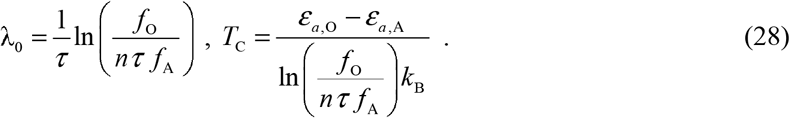

Note: Given that a single infected person spreads the disease to *n* ≈ 3 other persons per day for τ ≈ 5.2 days, with a deviation ±1 person, the propagated error is *δ T*_*c*_ = *T*_*c*_ ^2^ / [*nτ* (*ε* _a,o_ − *ε* _a,A_) / *k*_B_ ≈ 2.3K, which is consistent with the error in the measurements of *T*_C_.

Next, we derive the activation energies, characterizing the connection of the coronavirus with the receptor, *ε*_*a*, A_, as well as the competing reaction of the dissolution of the coronavirus, *ε* _*a*,O_. Given the value of λ_0_*T*_C_ (that is, the value of the parameter *p*_2_ in Eq.(4a)), we find the activation energy difference,

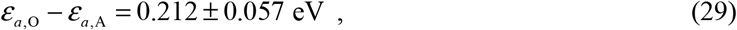

which coincides with the value derived experimentally in [32], i.e., eV. *ε*_*a*, O_ − *ε*_*a*, A_ = 5.35 kcal/mole = 0.232

Finally, we deal with the exact model of the temperature dependence, that is, we avoid the approximations of expanding the exponentials in Eq.(25) (though, we still consider *a*≈0, for simplicity). This is derived from combining Eq.(3), Eq.(23), and Eq.(25):

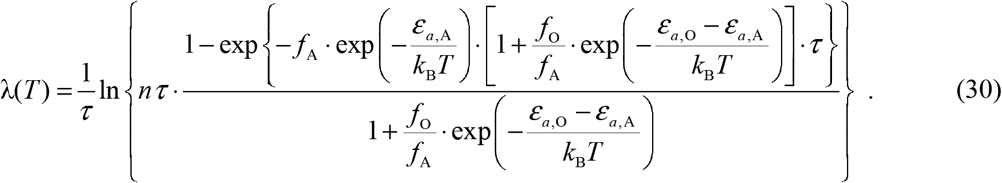

We have *ε*_*a*, O_ = 5.36 kcal/mole = 2697*k*_B_ [K], *ε*_*a*, O_ − *ε* _*a*,A_ = 5.35 kcal/mole = 2692*k*_B_ [K], τ = 5.2 d, while from *T*_C_∼303.25 ± 2.4 K (Eq.(5)), we derive:

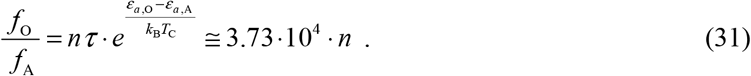

Hence, the model (30) becomes

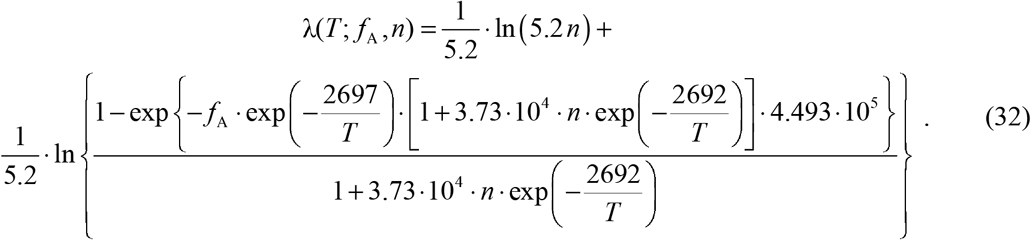

Figure 10 plots the model shown in Eq.(32), for fixed *n*=3d^-1^ (upper panels) and fixed frequency factor *f*_A_=0.01s^-1^ (lower panels).

**Figure 10.**
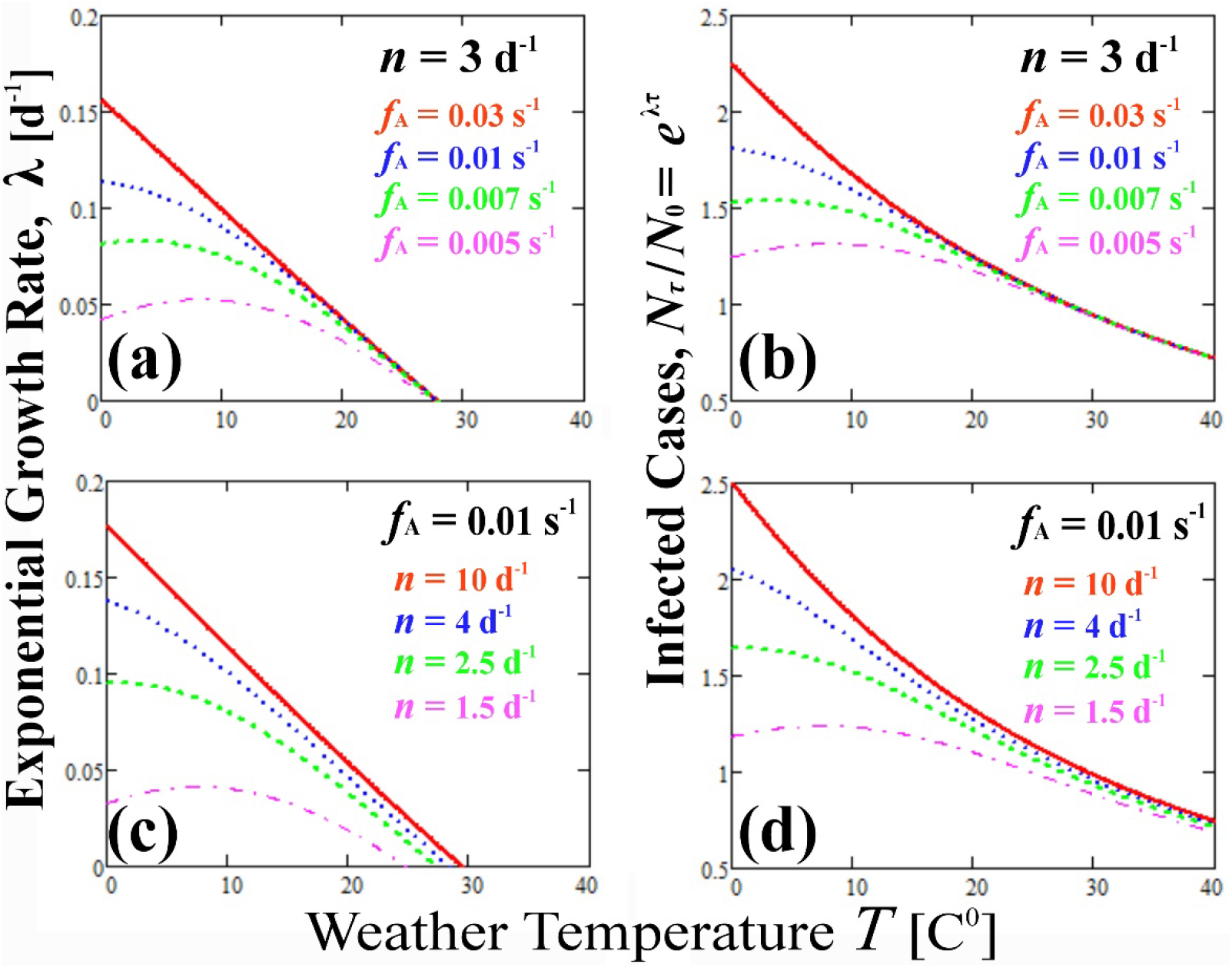
Exact model of the exponential growth rate (left panels) and the corresponding number of the infected cases in time *t*=τ (right panels), plotted with respect to temperature, for fixed *n*=3d^-1^ (upper panels) and fixed frequency factor *f*_A_=0.01s^-1^ (lower panels).

## 7. Discussion: What’s next?

It is straightforward to forecast the decline of the number of infected cases when the environmental temperature will climb above this critical value within 2021 for the Republic of Cyprus. The number of the daily new infected cases are expected to decline to zero after the 16^th^ of May, 2021, as shown in Figure 11.

**Figure 11.**
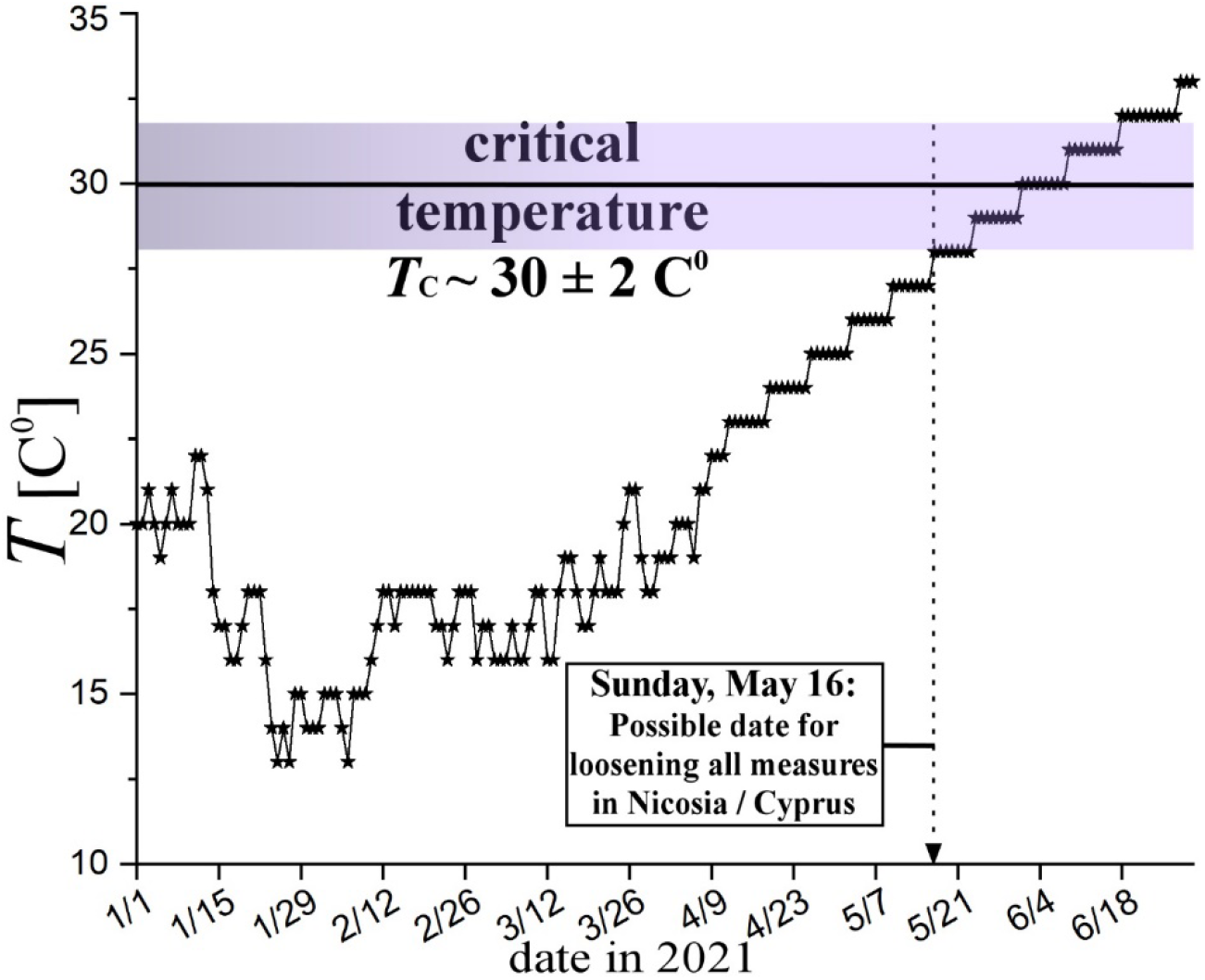
Plot of the average high temperature in Nicosia/Cyprus as recorded and forecasted for the dates in 2021. The temperature reaches, and climbs above, the critical temperature of *T*_C_ = 30 ± 2 C^0^ on the 16^th^ of May. Therefore, on this day forward, the number of cases infected by Covid-19 are expected to decline to zero. The plot suggests a possible date for loosening all the strict measures in the Republic of Cyprus, that is, the 16^th^ of May, 2021.

Encouraging as this result is, it is necessary to recall the conclusive statement from [1]. The resulting high statistical confidence of the negative correlation of the environmental temperature on the exponential growth rate of the cases infected by COVID-19 is certainly encouraging for loosening super-strict social-distancing measures, at least, during the summery high temperatures. However, we are, by no-means, recommending a return-to-work daily basis to be ensued by this analysis. Nonetheless, we do think that this work may be part of the decision, as well as an inspiration for repeating the same analysis in other heavily infected regions.

## 8. Conclusions

The paper improved our understanding of the effect and the functional dependence of the environmental temperature on the spread of COVID-19 and the rate of its exponential growth. We studied the daily number of the cases infected by Covid-19 during the second wave of the pandemic within 2020, as well as the daily number of the following deaths, and how they were affected by the daily average-high temperature for the districts of the Republic of Cyprus.

In particular, for the period from 8/25 to 12/31, that is, the second wave of the Covid-19 pandemic in 2020, we (i) examined the daily numbers of the infected cases with respect to the number and types of the tests performed, (ii) modelled the evolution of the number of the infected cases during the exponential growth phase, (iii) performed a statistical analysis for comparing the time series of the infected cases and the average-high environmental temperature, (iv) developed a model within the framework of chemical kinetics that describes the observed behavior and characteristics of the number of the infected cases and its exponential growth rate with respect to the environmental temperature, (v) performed a statistical analysis of the number of deaths caused by Covid-19 and their age distribution, and finally, (vi) understand the importance of the critical temperature that eliminates the exponential growth rate of the infected cases.

Among the main findings of the paper, are the following:

- Analyzed the results of the PCR and rapid antigen tests and showed that the average ratio of the PCR positive tests to the rapid positive tests is ∼ 2.57 ± 0.25; this coincides with the expected ratio of the PCR’s to the rapid test responses; the PCR’s response is characterized by a nonzero probability for more than 5 weeks and overestimates the positive tests (about 2.5 times, compared to the rapid test), and thus, the infected cases.
- Presented the common biases of the statistical sampling performed in the recording of the infected cases during the second wave of the Covid-19.
- Analyzed the time series of the age of deaths and showed that the average age shifts to older ages with rate ΔAge/Δ*t* ∼ 0.156 ± 0.050 [age year / day], i.e., about a year of age per week.
- Analyzed the total age distribution and showed that the probability of a person infected by Covid-19 to develop severe symptoms leading to death is strongly depended on the age of the person; in particular, the probability of having a death on the age of ∼67 or younger is less than 1/1000, for the second wave of the pandemic in Cyprus.
- Analyzed the data with respect to the environmental temperature and showed that the number of infected cases declined dramatically for temperatures reaching, and/or climbing above, the critical temperature of *T*_C_ = 30 ± 2 C^0^; since the number of the deaths is positively correlated to the number of the infected cases, similarly drop of the number of deaths is observed for high temperatures, above *T*_C_.
- Developed a model that describes the observed negative correlation between the exponential growth rate of the infected cases and the environmental temperature, which is based on the chemical kinetics of two parallel and competing reactions, describing the connection of the coronavirus towards the receptor, and the reaction of the dissolution of the coronavirus; the model results to the exact observed expression of the exponential rate with temperature.
- Estimated the difference of the activation energies corresponding to the chemical reactions of the connection of the coronavirus towards the receptor and of the coronavirus dissolution, i.e., *ε*_*a*,O_ − *ε*_*a*,A_ = 0.212 ± 0.057 eV, which is consistent with the experimentally derived value of *ε*_*a*,O_ − *ε*_*a*, A_ = 0.232 eV.
- Forecasted the decline of the number of infected cases when the environmental temperature climb above this critical value within the summery days of 2021 for the Republic of Cyprus; the daily new infected cases are expected to decline to zero after the 16^th^ of May, 2021.

## Data Availability

The sources of data used in the paper are publicly available.

